# Household income does not affect the pleiotropy of schizophrenia genetic liability with mental and physical health outcomes

**DOI:** 10.1101/2023.09.25.23296085

**Authors:** Manuela R. Kouakou, Brenda Cabrera-Mendoza, Gita A. Pathak, Tyrone D. Cannon, Renato Polimanti

## Abstract

**Background and Hypothesis:** Individuals with schizophrenia (SCZ) suffer from comorbidities that substantially reduce their life expectancy. Socioeconomic inequalities could contribute to many of the negative health outcomes associated with SCZ.

**Study Design:** We investigated genome-wide datasets related to SCZ (52,017 cases and 75,889 controls) from the Psychiatric Genomics Consortium, household income (HI; N=361,687) from UK Biobank, and 2,202 medical endpoints assessed in up to 342,499 FinnGen participants. A phenome-wide genetic correlation analysis of SCZ and HI was performed, also assessing whether SCZ genetic correlations were influenced by HI effect on SCZ. Additionally, SCZ and HI direct effects on medical endpoints were estimated using multivariable Mendelian randomization (MR).

**Study Results:** SCZ and HI showed overlapping genetic correlations with 70 traits (p<2.89×10^−5^), including mental health, substance use, gastrointestinal illnesses, reproductive outcomes, liver diseases, respiratory problems, and musculoskeletal phenotypes. SCZ genetic correlations with these traits were not affected by HI effect on SCZ. Considering Bonferroni multiple testing correction (p<7.14×10^−4^), MR analysis indicated that SCZ and HI may affect medical abortion (SCZ odds ratio, OR=1.07; HI OR=0.78), panic disorder (SCZ OR=1.20; HI OR=0.60), personality disorders (SCZ OR=1.31; HI OR=0.67), substance use (SCZ OR=1.2; HI OR=0.68), and adjustment disorders (SCZ OR=1.18; HI OR=0.78). Multivariable MR analysis confirmed that SCZ effects on these outcomes were independent of HI.

**Conclusions:** The effect of SCZ genetic liability on mental and physical health may not be strongly affected by socioeconomic differences. This suggests that SCZ-specific strategies are needed to reduce negative health outcomes affecting patients and high-risk individuals.

## INTRODUCTION

Individuals with schizophrenia (SCZ) are burdened with higher rates of physical and psychiatric illnesses than the general population^1^. These comorbidities contribute significantly to the excess mortality, with studies consistently reporting mortality rates two to three times higher than those of the general population ^2–4^. Several factors can be linked to poorer health outcomes and reduced life expectancy in individuals with SCZ. For instance, side effects of antipsychotic medications, suboptimal engagement in preventive care as well as unhealthy behaviors, such as substance use, lack of exercise, and unhealthy diet contribute to poorer overall health ^5, 6^.

Socioeconomic status (SES) may also play a pivotal role in adverse health outcomes among individuals with SCZ. In fact, individuals from lower socioeconomic backgrounds often face barriers to accessing healthcare services, including mental health care, resulting in delayed or inadequate interventions ^7^. Observational epidemiological studies have shown that negative health outcomes in SCZ patients are consistently associated with socioeconomic inequalities ^1^. However, since epidemiological studies can be affected by confounding and reverse causation, the underlying contribution of SES to these associations remains unclear.

Besides environmental factors, the complex genetic architecture of SCZ is also likely to contribute to the development of both psychiatric and somatic comorbidities. Indeed, several studies have shown a genetic overlap between SCZ and other psychiatric and somatic conditions ^8^. The association of genetic variants with human traits can be used to circumvent some of the biases that occur in epidemiological studies to generate more reliable evidence regarding the cause-effect relationships underlying SCZ comorbidities ^9^.

Recent studies demonstrated the importance of taking SES into account when investigating the pleiotropy of psychiatric disorders, behavioral traits, and other brain-related phenotypes^10^. In particular, genetically informed causal inference analyses showed how socioeconomic factors such as household income can partially explain certain associations observed in epidemiological studies ^11–14^. Leveraging data generated from genome-wide association studies (GWAS) conducted by the Psychiatric Genomics Consortium (PGC) ^15^ UK Biobank (UKB) ^16^ and FinnGen Project ^17^ we conducted a systematic investigation of whether economic factors may contribute at least partially to SCZ comorbidity with outcomes related to mental and physical health.

## METHODS

This study was performed leveraging genome-wide association statistics generated by previous studies. Because we used previously collected, de-identified data, this study did not require institutional review board approval. Applying different analytic approaches, we used genome-wide information to investigate the effect of socioeconomic factors on SCZ comorbidities. Specifically, we used genetic effects to explore the pleiotropy of SCZ and income on multiple health outcomes and then tested whether genetically inferred effects were independent of each other.

### Study Populations

#### Psychiatric Genomics Consortium

SCZ GWAS data were obtained from PGC meta-analysis ^18^. This included cohorts with different assessments. Cases included individuals affected by schizophrenia, schizoaffective disorder, or schizophrenia spectrum disorder (including non-affective psychosis). Controls included both individuals screened and unscreened for schizophrenia or other psychoses. Case definitions in PGC-SCZ cohorts were based on criteria from the diagnostic and statistical manual of mental disorders (DSM) and the international classification of diseases (ICD), diagnostic interviews, medical records, or hospital registers. In the present study, we used genome-wide association statistics generated from the GWAS meta-analysis of 76 cohorts including 52,017 SCZ cases and 75,889 controls.

#### UK Biobank

The UKB cohort includes approximately 500,000 participants aged between 40 and 69 recruited between 2006 and 2010 from across the United Kingdom. Participants provided samples for genetic analyses, as well as extensive information about their health, lifestyle, and medical history through questionnaires, interviews, and physical measurements ^16^. In the touchscreen questionnaire, UKB participants reported the average total household income before tax (hereafter referred to as income). This was assessed via a five-point scale: i) <£18 000; ii) £18 000–£30,999; iii) £31,000-£51,999; iv) £52,000-100,000; v) >£100,000. We obtained income genome-wide association statistics from the Pan-UKB analysis ^19^. This was performed using a generalized mixed model association testing framework available from the Scalable and Accurate Implementation of GEneralized (SAIGE) software ^20^. The following covariates were included in the Pan-UKB analysis: age, sex, ageXsex, age^2^, age^2^Xsex, and the top 10 within-ancestry principal components. Because PGC-SCZ GWAS was conducted in a sample of European descent, we investigated Pan-UKB income GWAS performed in 361,687 UKB participants of European descent.

#### FinnGen Project

The FinnGen Project enrolled a cohort of Finnish individuals, combining genomic data with comprehensive health information ^17^. Specifically, it integrates information from national health registries and electronic health records to derive data regarding diagnoses, medications, laboratory measurements, and other clinical outcomes. In the present study, we used the FinnGen Release DF8, which comprises genome-wide association statistics for 2,202 endpoints assessed in up to 342,499 participants of European descent (Supplemental Table 1). Association analysis was performed using regenie version 2.2.4 ^21^ using sex, age, top 10 within-ancestry principal components, and genotyping batch as covariates.

### Data Analyses

#### Genetic Correlation

SCZ and income genetic correlations with phenotypes available from UKB and FinnGen Project were computed using linkage disequilibrium (LD) score regression analysis v1.0.1 (LDSC) ^22^. Briefly, LDSC leverages LD information to estimate the proportion of heritability shared between two traits. Firstly, LD scores, which capture the average pairwise correlation of single nucleotide polymorphisms (SNPs) across the genome, were calculated for each SNP using the Phase 3 European populations from 1000 Genomes Project ^23^. Subsequently, these LD scores were used to estimate the heritability of each trait and finally, genetic correlations were computed using LD scores and heritability estimates for each trait. In line with LDSC guidelines, the analysis was limited to testing 1,217,311 SNPs present in the HapMap 3 reference panel ^24^. As recommended ^25^, the MHC region was excluded from the LDSC analysis, because of its complex LD pattern.

#### Multi-trait Conditioning

To assess the impact of income on SCZ, we used multi-trait conditioning and joint analysis (mtCOJO) ^26^, which is part of the GCTA (Genome-wide Complex Trait Analysis; Yang et al., 2011) software package. Briefly, conditional analysis involves examining the association between a genetic variant and a trait of interest while controlling for the effects of other traits. This method allows us to determine whether the association between the variant and the trait is independent of other correlated traits. By analyzing genome-wide association statistics for SCZ and income, mtCOJO enabled us to estimate the SCZ per-SNP effects after controlling for the effect of income on SCZ.

#### Mendelian Randomization

Mendelian randomization (MR) is a statistical method used to assess causal relationships between an exposure and an outcome of interest. It leverages genetic variants as instrumental variables to mimic a randomized controlled trial (RCT) setting, where the random allocation of genetic variants at conception is analogous to the random assignment of treatments in an RCT ^27^. In the present study, we applied the two-sample MR statistical method using the TwoSampleMR R package (v0.5.6) ^28^ to estimate the causal effect of SCZ and income on phenotypes that were identified as genetically correlated with both in the LDSC. Using the 1000 Genomes Project LD reference panel data ^23^, only LD-independent (r^2^=0.001 within a 10,000-kb window) genome-wide significant variants (P<5×10^−8^) were selected as genetic instruments. The effect size for the instruments on the exposures and the outcomes were then harmonized. We applied five MR methods, inverse-variance weighted (IVW), MR-Egger, weighted median, simple mode, weighted mode, and MR-RAPS (Robust Adjusted Profile Score) ^28, 29^. The outcome of the IVW method was considered as primary result due to its higher statistical power compared to the others ^28^. The other four methods were used to assess the concordance in the direction of effects. The instruments’ validity was also assessed by checking for horizontal pleiotropy, which occurs when the instrumental variables affect the outcome through pathways other than the exposure. Accordingly, we performed the IVW heterogeneity and MR-Egger intercept tests. Finally, MR-PRESSO (Mendelian Randomization Pleiotropy RESidual Sum and Outlier) method ^30^ was used to detect pleiotropic outliers and recalculate IVW estimates after their removal.

To assess whether SCZ and income effects are independent of each other, we performed multivariable MR (MVMR) ^31^. In line with the two-sample MR analysis, only LD-independent (r^2^=0.001 within a 10,000-kb window) genome-wide significant variants (P<5×10^−8^) were selected as genetic instruments from the SCZ and income GWAS results. The outliers identified in the previous MR-PRESSO analysis were excluded. The MVMR was performed using the R package MVMR ^32^.

## RESULTS

We tested the genetic correlation of SCZ and income with 1,732 heritable phenotypes available from FinnGen (Supplemental Table 2). After Bonferroni correction accounting for the number of traits tested, SCZ and income showed overlapping genetic correlations with a wide range of health outcomes assessed in the FinnGen cohort (Figure 1; N=70, p<2.89×10^-5^). The pattern was generally in line with the negative genetic correlation between SCZ and income (rg= -0.14, p=4.71×10^−12^). The health outcomes included mental health (e.g., personality disorders: SCZ rg=0.53, p=2.89×10^−50^; income rg=-0.32, p=3.19×10^−19^), substance use (e.g., smoking: SCZ rg=0.24, p=3.36×10^−6^; income rg=-0.62, p=4.4×10^−19^), gastrointestinal illnesses (e.g., other diseases of the digestive system: SCZ rg=0.31, p=2.02×10^−5^; income rg=-0.41, p=8.17×10^−7^), reproductive outcomes (e.g., medical abortion: SCZ rg=0.27, p=7.26×10^−18^; income rg=-0.49, p=1.54×10^−45^), oral health (e.g., gingivitis and periodontal diseases: SCZ rg=0.22, p=4.85×10^−7^; income rg=-0.21, p=1.66×10^−6^), blood-related conditions (e.g., anemia: SCZ rg=0.20, p=1.02×10^−7^; income rg=-0.36, p=5.14×10^−16^), liver diseases (e.g., viral hepatitis: SCZ rg=0.34, p=9.82×10^−7^; income rg=-0.40, p=6.54×10^-8^), respiratory problems (e.g., early-onset chronic obstructive pulmonary disease: SCZ rg=0.14, p=8.5×10^-6^; income rg=-0.41, p=1.54×10^−23^), and mortality (any cause of death: SCZ rg=0.19, p=3.78×10^-6^; income rg=-0.46, p=4.28×10^−15^). Differently from the overall patterns (i.e., positive genetic correlation with SCZ and negative genetic correlation with income), we also identified eleven musculoskeletal phenotypes showing negative genetic correlations with both SCZ and income (e.g., disorders of synovium and tendon: SCZ rg=- 0.18, p=6.27×10^-6^; income rg=-0.33, p=5.23×10^−18^).

**Figure 1:**
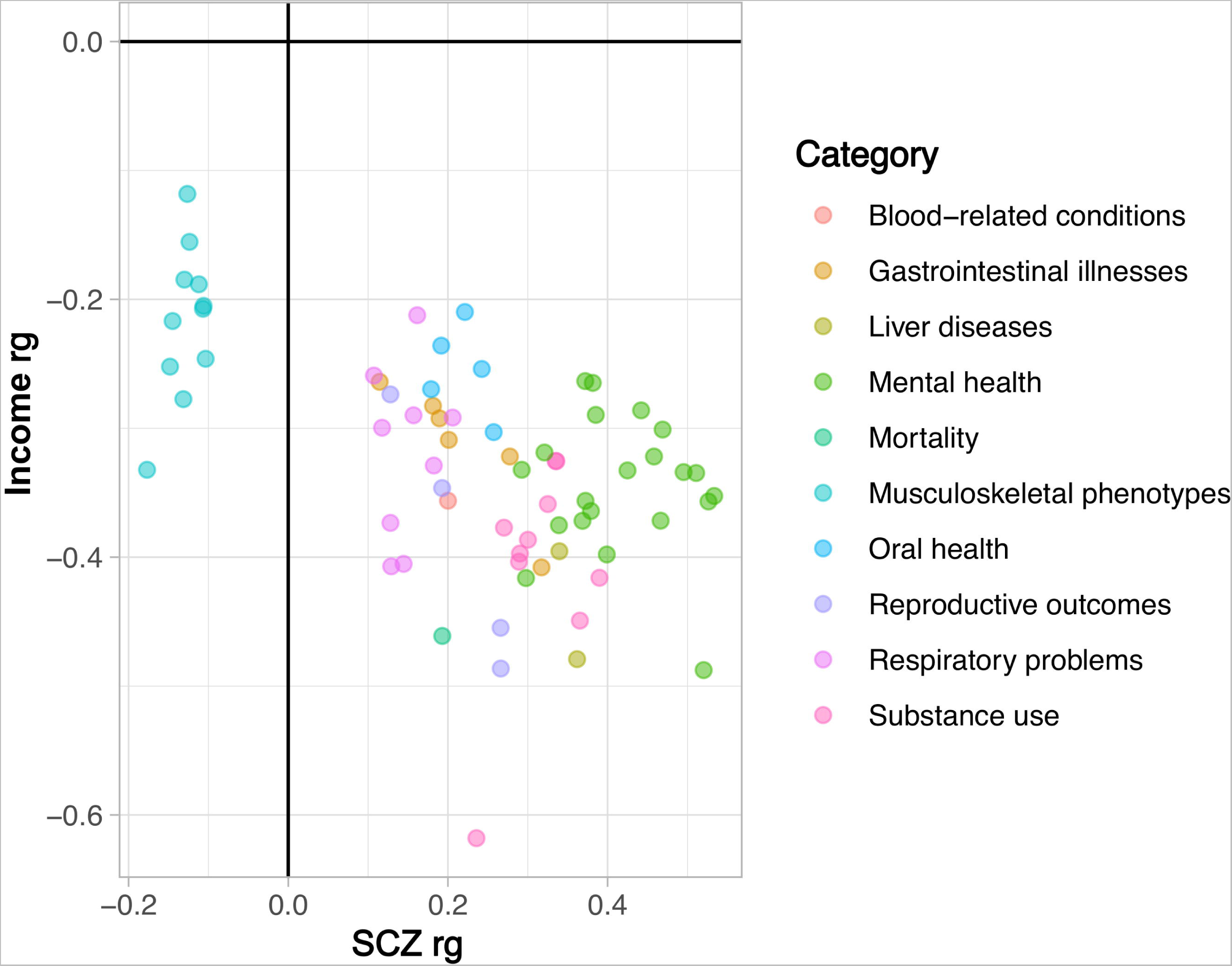
Genetic correlation of schizophrenia (SCZ) and household income (Income) with medical endpoints after Bonferroni multiple testing correction (p<2.89×10^-5^). Full results are available in Supplemental Table 2.

To verify whether these genetic correlations were influenced by the effect of income on SCZ, we used the mtCOJO approach ^26^. Specifically, we estimated SCZ per-SNP effect controlling for the putative causal effect of income on SCZ (odds ratio, OR=0.93, 95% confidence interval, CI=0.87-0.99). After income conditioning of genome-wide association statistics of SCZ, we recalculated SCZ genetic correlation with the phenotypes available from FinnGen. Comparing SCZ genetic correlation before and after income conditioning, we did not observe any statistical difference in the rg estimates (Supplemental Table 3).

Although income did not affect SCZ pleiotropy with health outcomes, this does not exclude that income may at least partially contribute to some of SCZ comorbidities. To test this hypothesis, we applied the MVMR framework, which is similar to a factorial randomized trial where multiple treatments are tested (Burgess et al., 2015). Initially, we performed a two-sample MR analysis testing the effect of SCZ and income on genetically correlated traits separately. After removing outliers identified by the MR-PRESSO analysis and excluding results affected by significant heterogeneity and horizontal pleiotropy, SCZ and income showed possible causal effects on five phenotypes after Bonferroni multiple testing correction (Table 1; Supplemental Tables 4 and 5): medical abortion (SCZ: OR=1.07, 95% CI=1.03-1.11; income OR=0.78, 95% CI=0.70-0.87), panic disorder (SCZ OR=1.20, 95% CI=1.13-1.29; income OR=0.6, 95% CI=0.48-0.76), personality disorders (SCZ OR=1.31, 95% CI=1.24-1.4; income OR=0.67, 95% CI=0.55-0.82), substance use excluding alcohol (SCZ OR=1.2, 95% CI=1.13-1.27; income OR=0.67, 95% CI=0.55-0.85); adjustment disorders (SCZ OR=1.18, 95% CI=1.13-1.23; income OR=0.78, 95% CI=0.68-0.90).

**Table 1.**
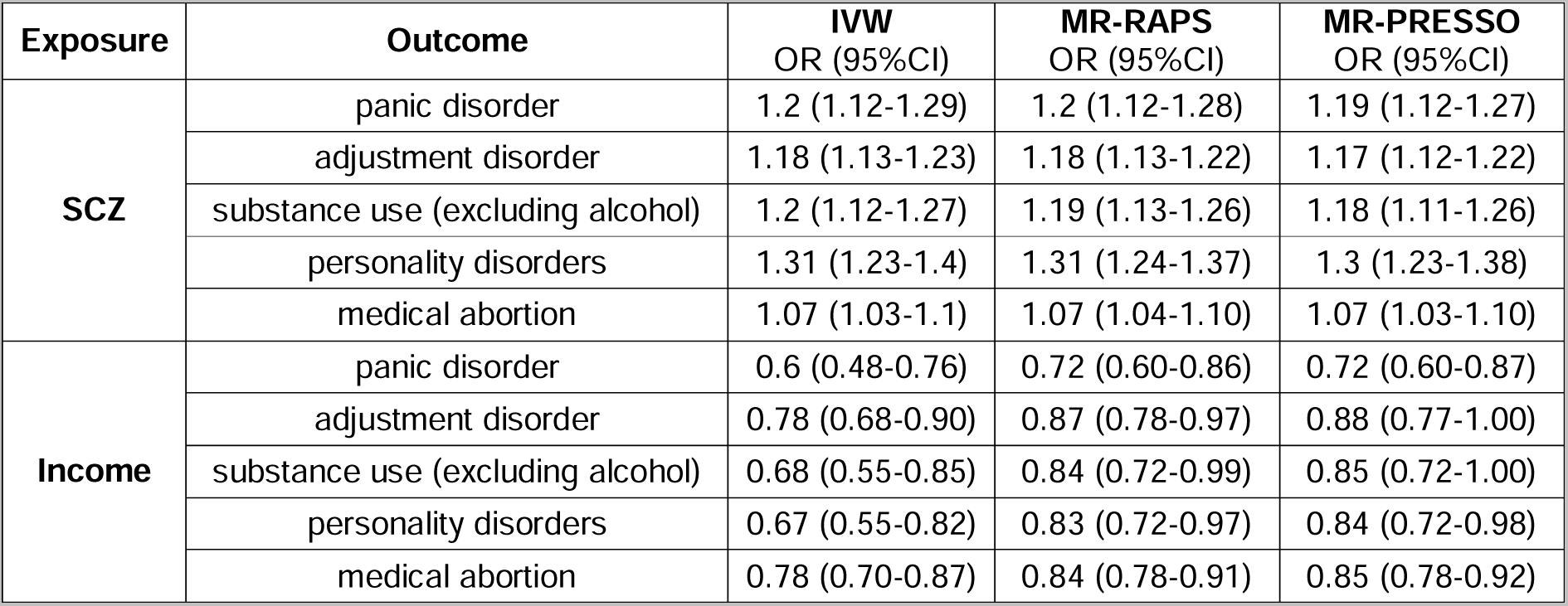
Odds ratios (OR) and 95% confidence intervals (95%CI) related to the effect of schizophrenia genetic liability (SCZ) and household income (income) on medical endpoints surviving Bonferroni multiple testing correction and sensitivity analyses. The estimates reported were obtained using inverse-variance weighted (IVW), MR- Robust Adjusted Profile Score (MR-RAPS), and MR-PRESSO (Mendelian Randomization Pleiotropy RESidual Sum and Outlier). Full Results are available in Supplemental Tables 4 and 5.

MVMR analysis showed that the effect of SCZ genetic liability on these endpoints is independent of income (Figure 2): OR_MVMR_=1.05 (95%CI_MVMR_=1.01-1.09) for medical abortion; OR_MVMR_=1.2 (95%CI_MVMR_=1.11-1.3) for panic disorder; OR_MVMR_=1.34 (95%CI_MVMR_=1.25-1.43) for personality disorders; OR_MVMR_=1.17 (95%CI_MVMR_=1.09-1.25) for substance use excluding alcohol; OR_MVMR_=1.15 (95%CI_MVMR_= 1.1-1.22) for adjustment disorders.

**Figure 2:**
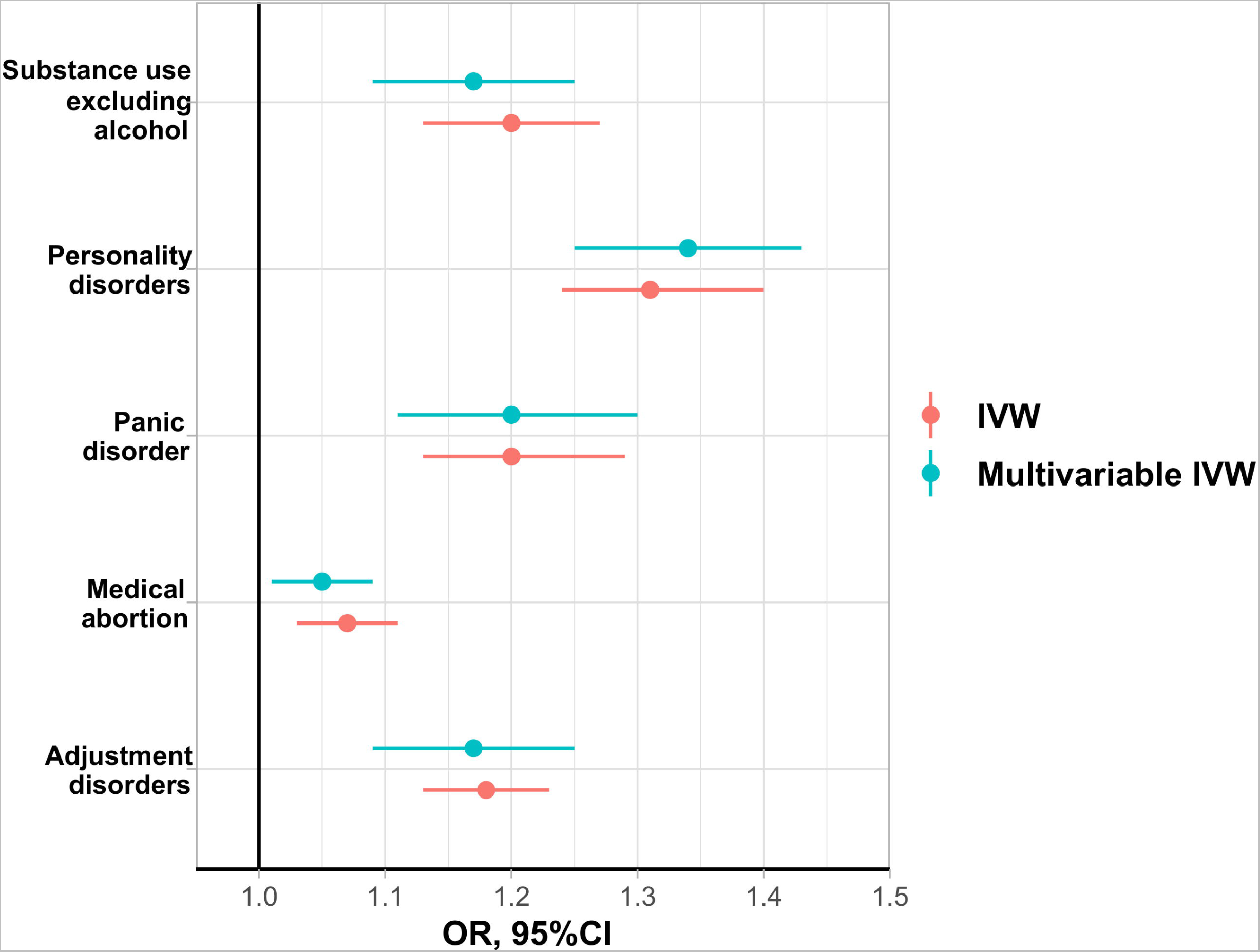
Association of schizophrenia genetic liability with medical endpoints estimated using the inverse-variance weighted (IVW) approach considering univariable and multivariable Mendelian randomization (MR) models. Estimates are reported as odds ratios (OR) and 95% confidence intervals (95%CI). The multivariable MR model accounted for the effect of household income on the endpoints tested.

## DISCUSSION

The present study provided evidence that the pleiotropy of SCZ genetic liability with mental and physical health appears to be independent of the effect of income. Overall, SCZ and income were genetically correlated with psychiatric disorders, substance use, personality traits as well as a range of somatic traits and diseases. These included several negative outcomes known to contribute to the increased morbidity and mortality observed among SCZ patients such as respiratory problems and gastrointestinal illnesses ^33^. In line with the negative genetic overlap between SCZ and income, the majority of the medical endpoints tested showed genetic correlations with opposite direction: positive genetic correlation with SCZ and negative genetic correlation with income. However, in contrast with this pattern, multiple musculoskeletal phenotypes investigated in our study showed a negative genetic correlation with both SCZ and income. While a lower prevalence of rheumatoid arthritis (RA) in SCZ patients ^34, 35^ and negative genetic overlap between the two conditions due to the MHC region has been previously reported ^36, 37^, the epidemiological evidence for the direction of correlation with non-inflammatory joint disorder is inconsistent ^38, 39^. It has been hypothesized that SCZ patients may manifest fewer joint pain symptoms due to low sensitivity to chronic pain ^40^. Because we excluded the MHC region from the LDSC analysis, SCZ genetic correlations with musculoskeletal phenotypes reported in the present study are due to the contribution of many variants across the genome and not only to the effect of the MHC region. Further analyses of the genome-wide pleiotropy between SCZ and musculoskeletal phenotypes could permit to disentangle whether this is due to the effect of pain sensitivity or to pathogenic pathways that affect the brain leading to increased SCZ risk and peripheral tissues leading to reduced musculoskeletal outcomes.

Similar to other social determinants of health, economic inequality has been consistently associated with SCZ risk, treatment effectiveness, and negative outcomes in SCZ patients ^41,42^. Furthermore, previous studies reported a significant difference in heritability as well as genetic correlations between psychiatric traits when income and other SES factors were taken into account ^10, 43^. To investigate whether income at least partially explained SCZ genetic overlap with mental and physical outcomes, we performed a conditional analysis to estimate SCZ genetic correlation with medical endpoints accounting for the effect of income on SCZ. Our results showed that controlling for the impact of income on SCZ did not change SCZ genetic correlation estimates. This supports that economic factors might not affect SCZ pleiotropy with health outcomes. However, the findings of our conditional analyses do not exclude that income may at least partially contribute to some SCZ comorbidities. To test this hypothesis, we applied a MR design, identifying five medical endpoints with a possible causal relationship with both SCZ and income. These included four phenotypes related to mental health (i.e., panic disorder, adjustment disorder, substance use excluding alcohol, and personality disorders) and one related to reproductive health (i.e., medical abortion). Consistent with the conditional analysis of SCZ genetic correlations, MVMR approach provided further evidence that SCZ effect on these traits is independent of income. The independence of SCZ genetic liability and income suggests possible additive effects of these factors with respect to the five medical endpoints identified where individuals with high SCZ risk may experience an additional effect due to economic inequalities. For example, women affected by SCZ are well-known to have worse sexual and reproductive health with higher risk of coerced sex, sexual risk behavior, unwanted pregnancies, abortion, and obstetric complications ^44^. In our study, we show that SCZ genetic liability and income may have independent effects on medical abortion. Epidemiological studies showed that women with SCZ have higher rates of medical abortions than women without SCZ ^45^. The relationship between economic inequality and medical abortion is strongly affected by sociocultural context, where factors like abortion legal status and stigma also play major roles ^46, 47^. Indeed, economic, legal, social, and cultural factors all contribute to generate differences among women in cognitive and material resources needed when making a decision regarding performing a medical abortion or continuing the pregnancy. While previous studies showed that SCZ is associated with medical abortion also when controlling for income and other factors related to material deprivation ^48^, our genetically informed causal inference analysis highlights that SCZ association with medical abortion is independent of the effects of economic inequalities on SCZ and on medical abortion itself. This supports that further studies should focus on identifying which SCZ-specific factors contribute to the increased medical abortions observed among women affected by SCZ.

In line with SCZ psychiatric comorbidities and the impact of economic factors on mental health, we identified multiple causal relationships linking SCZ and income with a range of mental illnesses. Among them, personality disorders appear to be strongly affected by both SCZ genetic liability and income. Historically, personality changes were seen as part of SCZ pathogenesis, being also considered evidence of its premorbid predisposition ^49^. In recent years, clinicians clarified the importance of differentiating among the diagnoses of SCZ, psychosis-prone schizotypal personality disorder, and borderline personality disorder ^50^. However, because of the comorbidity between SCZ and personality disorders, a thorough personality assessment has been proposed as an integral part of a SCZ assessment ^50^. This is supported by the effect of SCZ genetic liability on personality disorders observed in the present study. Specifically, the fact that this association is independent of income supports that the relationship between SCZ and personality disorders is likely due to shared pathogenic mechanisms rather than negative environmental factors linked to economic inequalities. A similar scenario can also explain the effect of SCZ genetic liability on substance use. Indeed, SCZ is often found comorbid with substance use, a phenomenon known as dual diagnosis ^51^. Multiple MR studies reported SCZ associations with cannabis and tobacco use and misuse ^52–55^. In our study, we demonstrate that this relationship appears to be independent of economic factors, supporting a direct role of SCZ genetic liability in the predisposition to substance use. We observed similar evidence also for SCZ associations with panic disorder and adjustment disorder. Previous polygenic risk score investigations and MR analyses showed genetic overlap and possible cause effects linking SCZ to anxiety disorders, including panic disorder ^56, 57^. Although economic factors have a major impact on panic disorders and other anxiety-related outcomes ^58^, our genetically informed analyses support a direct relationship of SCZ genetic liability with panic disorder, which appears to be independent of an income effect. With respect to adjustment disorder, previous MR analyses showed the important role of economic factors in the context of comorbidity of trauma-related outcomes such as posttraumatic stress disorder ^11, 13^. However, the present study supports that economic factors should not play a role in the dynamics responsible for SCZ comorbidity with adjustment disorder. Further studies will be needed to confirm this finding and verify whether this applies also to other trauma-related outcomes.

Our study has some limitations. First, income may not comprehensively capture the complexity of socioeconomic factors, which also include material deprivation and educational attainment. A genetic study showed that the cumulative effect of different SES measures is larger than the individual effect of income alone although the effects among income, deprivation index, and educational attainment were largely consistent ^43^. Second, differences in the effective sample size among the FinnGen endpoints investigated may have impacted our statistical power to investigate health outcomes with low prevalence in the general population. Third, although we identified a female-specific association (medical abortion), the lack of sex-stratified SCZ genome-wide association statistics did not permit us to fully explore possible female and male-specific relationships. Finally, our analysis was based on data from individuals of European descent due to the lack of large-scale GWAS available for other ancestry groups. It will be important to confirm and expand our findings in other population groups because of the known racial disparities in SCZ ^59^.

In summary, we performed a comprehensive analysis of genetic overlap and possible causal effects contributing to SCZ comorbidities, observing that economic factors may play an independent role in the genetically inferred associations observed. This supports that SCZ-specific factors, rather than economic factors per se, are likely to contribute to the comorbidities observed among the affected patients. Further studies will be needed to explore these SCZ-specific mechanisms, with the aim of tailoring interventions, allocating resources effectively, and improving health outcomes for patients and individuals at high risk.

## Data Availability

All data produced in the present work are contained in the manuscript.

## ACKNOWLEDGEMENTS

The authors thank participants and investigators involved in the Psychiatric Genomics Consortium, the UK Biobank, and the FinnGen Project.

## REFERENCES

1. Burns JK, Tomita A, Kapadia AS. Income inequality and schizophrenia: increased schizophrenia incidence in countries with high levels of income inequality. Int J Soc Psychiatry Mar 2014;60(2):185–196.

2. Kugathasan P, Stubbs B, Aagaard J, Jensen SE, Munk Laursen T, Nielsen RE. Increased mortality from somatic multimorbidity in patients with schizophrenia: a Danish nationwide cohort study. Acta Psychiatr Scand Oct 2019;140(4):340–348.

3. Brown S. Excess mortality of schizophrenia: A meta-analysis. The British Journal of Psychiatry 1997;171(6):502–508.

4. Saha S, Chant D, McGrath J. A Systematic Review of Mortality in Schizophrenia: Is the Differential Mortality Gap Worsening Over Time? Archives of General Psychiatry 2007;64(10):1123–1131.

5. Correll CU, Detraux J, De Lepeleire J, De Hert M. Effects of antipsychotics, antidepressants and mood stabilizers on risk for physical diseases in people with schizophrenia, depression and bipolar disorder. World Psychiatry Jun 2015;14(2):119–136.

6. Kerfoot KE, Rosenheck RA, Petrakis IL, Swartz MS, Keefe RS, McEvoy JP, Stroup TS, Investigators C. Substance use and schizophrenia: adverse correlates in the CATIE study sample. Schizophr Res Nov 2011;132(2-3):177–182.

7. Adler NE, Newman K. Socioeconomic disparities in health: pathways and policies. Health Aff (Millwood) Mar-Apr 2002;21(2):60–76.

8. Smeland OB, Frei O, Dale AM, Andreassen OA. The polygenic architecture of schizophrenia - rethinking pathogenesis and nosology. Nat Rev Neurol Jul 2020;16(7):366–379.

9. Davey Smith G, Hemani G. Mendelian randomization: genetic anchors for causal inference in epidemiological studies. Hum Mol Genet Sep 15 2014;23(R1):R89–98.

10. Wendt FR, Pathak GA, Lencz T, Krystal JH, Gelernter J, Polimanti R. Multivariate genome-wide analysis of education, socioeconomic status and brain phenome. Nat Hum Behav Apr 2021;5(4):482–496.

11. Polimanti R, Ratanatharathorn A, Maihofer AX, et al. Association of Economic Status and Educational Attainment With Posttraumatic Stress Disorder: A Mendelian Randomization Study. JAMA Netw Open May 3 2019;2(5):e193447.

12. Cabrera-Mendoza B, Wendt FR, Pathak GA, De Angelis F, De Lillo A, Koller D, Polimanti R. The association of obesity-related traits on COVID-19 severity and hospitalization is affected by socio-economic status: a multivariable Mendelian randomization study. Int J Epidemiol Oct 13 2022;51(5):1371–1383.

13. Muniz Carvalho C, Wendt FR, Maihofer AX, et al. Dissecting the genetic association of C-reactive protein with PTSD, traumatic events, and social support. Neuropsychopharmacology May 2021;46(6):1071–1077.

14. Tylee DS, Lee YK, Wendt FR, Pathak GA, Levey DF, De Angelis F, Gelernter J, Polimanti R. An Atlas of Genetic Correlations and Genetically Informed Associations Linking Psychiatric and Immune-Related Phenotypes. JAMA Psychiatry Jul 1 2022;79(7):667–676.

15. Sullivan PF, Agrawal A, Bulik CM, et al. Psychiatric Genomics: An Update and an Agenda. Am J Psychiatry Jan 1 2018;175(1):15–27.

16. Bycroft C, Freeman C, Petkova D, et al. The UK Biobank resource with deep phenotyping and genomic data. Nature Oct 2018;562(7726):203-209.

17. Kurki MI, Karjalainen J, Palta P, et al. FinnGen provides genetic insights from a well-phenotyped isolated population. Nature Jan 2023;613(7944):508-518.

18. Trubetskoy V, Pardiñas AF, Qi T, et al. Mapping genomic loci implicates genes and synaptic biology in schizophrenia. Nature Apr 2022;604(7906):502-508.

19. team P-U. Available at: https://pan.ukbb.broadinstitute.org.

20. Zhou W, Nielsen JB, Fritsche LG, et al. Efficiently controlling for case-control imbalance and sample relatedness in large-scale genetic association studies. Nat Genet Sep 2018;50(9):1335–1341.

21. Mbatchou J, Barnard L, Backman J, et al. Computationally efficient whole-genome regression for quantitative and binary traits. Nature Genetics 2021/07/01 2021;53(7):1097-1103.

22. Bulik-Sullivan BK, Loh P-R, Finucane HK, et al. LD Score regression distinguishes confounding from polygenicity in genome-wide association studies. Nature Genetics 2015/03/01 2015;47(3):291–295.

23. Genomes Project C, Auton A, Brooks LD, et al. A global reference for human genetic variation. Nature Oct 1 2015;526(7571):68-74.

24. International HapMap C, Altshuler DM, Gibbs RA, et al. Integrating common and rare genetic variation in diverse human populations. Nature Sep 2 2010;467(7311):52-58.

25. Finucane HK, Bulik-Sullivan B, Gusev A, et al. Partitioning heritability by functional annotation using genome-wide association summary statistics. Nat Genet Nov 2015;47(11):1228–1235.

26. Zhu Z, Zheng Z, Zhang F, et al. Causal associations between risk factors and common diseases inferred from GWAS summary data. Nature Communications 2018/01/15 2018;9(1):224.

27. Ference BA, Holmes MV, Smith GD. Using Mendelian Randomization to Improve the Design of Randomized Trials. Cold Spring Harb Perspect Med Jul 1 2021;11(7).

28. Hemani G, Zheng J, Elsworth B, et al. The MR-Base platform supports systematic causal inference across the human phenome. Elife May 30 2018;7.

29. Zhao Q, Wang J, Hemani G, Bowden J, Small DS. Statistical inference in two-sample summary-data Mendelian randomization using robust adjusted profile score. The Annals of Statistics 2020;48(3):1742–1769, 1728.

30. Verbanck M, Chen CY, Neale B, Do R. Detection of widespread horizontal pleiotropy in causal relationships inferred from Mendelian randomization between complex traits and diseases. Nat Genet May 2018;50(5):693–698.

31. Sanderson E, Davey Smith G, Windmeijer F, Bowden J. An examination of multivariable Mendelian randomization in the single-sample and two-sample summary data settings. Int J Epidemiol Jun 1 2019;48(3):713–727.

32. Yavorska OO, Burgess S. MendelianRandomization: an R package for performing Mendelian randomization analyses using summarized data. International Journal of Epidemiology 2017;46(6):1734–1739.

33. Correll CU, Solmi M, Croatto G, et al. Mortality in people with schizophrenia: a systematic review and meta-analysis of relative risk and aggravating or attenuating factors. World Psychiatry Jun 2022;21(2):248–271.

34. Chen SJ, Chao YL, Chen CY, et al. Prevalence of autoimmune diseases in in-patients with schizophrenia: nationwide population-based study. Br J Psychiatry May 2012;200(5):374–380.

35. Oken RJ, Schulzer M. At Issue: Schizophrenia and Rheumatoid Arthritis: The Negative Association Revisited. Schizophrenia Bulletin 1999;25(4):625–638.

36. Zamanpoor M, Ghaedi H, Omrani MD. The genetic basis for the inverse relationship between rheumatoid arthritis and schizophrenia. Mol Genet Genomic Med Nov 2020;8(11):e1483.

37. Sellgren C, Frisell T, Lichtenstein P, Landèn M, Askling J. The association between schizophrenia and rheumatoid arthritis: a nationwide population-based Swedish study on intraindividual and familial risks. Schizophr Bull Nov 2014;40(6):1552–1559.

38. Tyrovolas S, Moneta V, Giné Vázquez I, Koyanagi A, Abduljabbar AS, Haro JM. Mental Disorders, Musculoskeletal Disorders and Income-Driven Patterns: Evidence from the Global Burden of Disease Study 2017. J Clin Med Jul 10 2020;9(7).

39. Huang SW, Wang WT, Lin LF, Liao CD, Liou TH, Lin HW. Association between psychiatric disorders and osteoarthritis: a nationwide longitudinal population-based study. Medicine (Baltimore) Jun 2016;95(26):e4016.

40. Lévesque M, Potvin S, Marchand S, Stip E, Grignon S, Pierre L, Lipp O, Goffaux P. Pain perception in schizophrenia: evidence of a specific pain response profile. Pain Med Dec 2012;13(12):1571–1579.

41. Bennett D, Rosenheck R. Socioeconomic status and the effectiveness of treatment for first-episode psychosis. Health Serv Res Jun 2021;56(3):409–417.

42. Jester DJ, Thomas ML, Sturm ET, et al. Review of Major Social Determinants of Health in Schizophrenia-Spectrum Psychotic Disorders: I. Clinical Outcomes. Schizophr Bull Jul 4 2023;49(4):837–850.

43. Marees AT, Smit DJA, Abdellaoui A, et al. Genetic correlates of socio-economic status influence the pattern of shared heritability across mental health traits. Nat Hum Behav Aug 2021;5(8):1065–1073.

44. Barker LC, Vigod SN. Sexual health of women with schizophrenia: A review. Front Neuroendocrinol Apr 2020;57:100840.

45. Brown HK, Dennis C-L, Kurdyak P, Vigod SN. A population-based study of the frequency and predictors of induced abortion among women with schizophrenia. The British Journal of Psychiatry 2019;215(6):736–743.

46. Bearak J, Popinchalk A, Ganatra B, Moller AB, Tunçalp Ö, Beavin C, Kwok L, Alkema L. Unintended pregnancy and abortion by income, region, and the legal status of abortion: estimates from a comprehensive model for 1990-2019. Lancet Glob Health Sep 2020;8(9):e1152–e1161.

47. Sorhaindo AM, Lavelanet AF. Why does abortion stigma matter? A scoping review and hybrid analysis of qualitative evidence illustrating the role of stigma in the quality of abortion care. Soc Sci Med Aug 24 2022;311:115271.

48. Fabre C, Pauly V, Baumstarck K, et al. Pregnancy, delivery and neonatal complications in women with schizophrenia: a national population-based cohort study. Lancet Reg Health Eur Nov 2021;10:100209.

49. Kraepelin E. Dementia praecox and paraphrenia, 2002.

50. Simonsen E, Newton-Howes G. Personality Pathology and Schizophrenia. Schizophr Bull Oct 17 2018;44(6):1180–1184.

51. Polimanti R, Agrawal A, Gelernter J. Schizophrenia and substance use comorbidity: a genome-wide perspective. Genome Medicine 2017/03/21 2017;9(1):25.

52. Gage SH, Jones HJ, Burgess S, Bowden J, Davey Smith G, Zammit S, Munafò MR. Assessing causality in associations between cannabis use and schizophrenia risk: a two-sample Mendelian randomization study. Psychol Med Apr 2017;47(5):971–980.

53. Pasman JA, Verweij KJH, Gerring Z, et al. GWAS of lifetime cannabis use reveals new risk loci, genetic overlap with psychiatric traits, and a causal influence of schizophrenia. Nat Neurosci Sep 2018;21(9):1161–1170.

54. Wootton RE, Richmond RC, Stuijfzand BG, et al. Evidence for causal effects of lifetime smoking on risk for depression and schizophrenia: a Mendelian randomisation study. Psychol Med Oct 2020;50(14):2435–2443.

55. Gage SH, Jones HJ, Taylor AE, Burgess S, Zammit S, Munafò MR. Investigating causality in associations between smoking initiation and schizophrenia using Mendelian randomization. Sci Rep Jan 19 2017;7:40653.

56. Jones HJ, Martin D, Lewis SJ, Davey Smith G, O’Donovan MC, Owen MJ, Walters JTR, Zammit S. A Mendelian randomization study of the causal association between anxiety phenotypes and schizophrenia. Am J Med Genet B Neuropsychiatr Genet Sep 2020;183(6):360–369.

57. Richards A, Horwood J, Boden J, et al. Associations between schizophrenia genetic risk, anxiety disorders and manic/hypomanic episode in a longitudinal population cohort study. The British Journal of Psychiatry 2019;214(2):96–102.

58. de Jonge P, Roest AM, Lim CC, et al. Cross-national epidemiology of panic disorder and panic attacks in the world mental health surveys. Depress Anxiety Dec 2016;33(12):1155–1177.

59. Schwartz RC, Blankenship DM. Racial disparities in psychotic disorder diagnosis: A review of empirical literature. World J Psychiatry Dec 22 2014;4(4):133–140.

